# Estimating the Case Fatality Risk of COVID-19 using Cases from Outside China

**DOI:** 10.1101/2020.02.15.20023499

**Authors:** Nick Wilson, Amanda Kvalsvig, Lucy Telfar Barnard, Michael G Baker

**Author notes:** Correspondence: Prof Nick Wilson.

## Abstract

There is large uncertainty around the case fatality risk (CFR) for COVID-19 in China. Therefore, we considered symptomatic cases outside of China (countries/settings with 20+ cases) and the proportion who are in intensive care units (4.0%, 14/349 on 13 February 2020). Given what is known about CFRs for ICU patients with severe respiratory conditions from a meta-analysis, we estimated a CFR of 1.37% (95%CI: 0.57% to 3.22%) for COVID- 19 cases outside of China.

---

The new coronavirus (COVID-19) appears to be fairly transmissible [1], and is spreading in China. Disease severity is a particularly important parameter for understanding this new disease [1], but unfortunately, the case fatality risk (CFR) data from China is difficult to interpret owing to likely missed mild cases (including due to a lack of appropriate test kits early in the epidemic) and also the likely delay in deaths occurring. Such a prolonged time course is suggested by a case series of 138 cases from Wuhan, China with cases enrolled between 1-28 January 2020. For this group on 3 February, 62% were still hospitalised and 31% (11/36) of those admitted to ICU were still there (6 had died) [2].

In jurisdictions outside China (and excluding Hong Kong, Macao and Taiwan) the CFR as detailed in the 13 February WHO Report [3] was 1/447 = 0.22% (95% confidence interval (CI) = 0.40% to 1.26%). But this estimate also did not account for possible missed mild cases in those countries or the lag time between hospitalisation and death.

One possible way to get a better estimate of the CFR is to consider the proportion of cases currently in intensive care units (ICUs) in settings outside China where the healthcare system is working relatively normally (ie, is not over-burdened by the epidemic). As per Table 1 we estimated that 4.0% (14/349) of identified cases were in ICUs in all the countries outside of China that had 20+ cases.

**Table 1.** Cases in countries and settings outside China with 20+ cases on 13 February 2020 and the proportion estimated to be in intensive care units

| Country/<br>setting | Confirmed<br>cases as of<br>13 February<br>[3] | Estimated<br>proportion<br>in ICU | Comments and references for ICU occupancy |
| --- | --- | --- | --- |
| Singapore | 50 | 16% (8/50) | Based on a news report on 12 February [4]. |
| Thailand | 33 | 1/33<br>(assumed as<br>per "serious<br>condition") | Based on a news report on 13 February [5]: "Confirmed cases reached 33, including one person in serious condition, by 12 February". |
| Japan | 29 | 0/25 | Based on a translation of a Ministry of Health, Labour and Welfare document on 12 February [6]. See below for separate cruise ship data. |
| Republic of<br>Korea | 28 | 0/23 (see<br>comments) | When there were 23 known cases: "all 22 patients currently isolated are in stable condition."... "one patient was discharged from a hospital yesterday after he was cured". Based on a news report on 6 February [7]. |
| International conveyance -<br>(specifically the cruise ship<br>"Diamond Princess" moored in Japan) | 174 (updated to 218 later on 13 February - see comments) | 5/218 (see comments) | Based on a news report on 13 February [8]: "Kato said five people from the ship are currently in serious condition in hospital". A prior report on 12 February in the same news source gave more detail for the first four people where the Health Minister (Katsunobu Kato) was reported saying: "At this point, we have confirmed that four people, among those who are hospitalised, are in a serious condition, either on a ventilator or in an intensive care unit." [9] |
| **Total** |  | **4.0%**<br>**(14/349)** |  |

Next we considered the typical survival of people admitted to ICUs with severe respiratory conditions. We used data from a meta-analysis of trials comparing higher vs lower levels of positive end-expiratory pressure (PEEP) in adults with acute lung injury or acute respiratory distress syndrome (ARDS) [10]. This study found 374 hospital deaths in 1136 patients (32.9%) assigned to treatment with higher PEEP and 409 hospital deaths in 1163 patients (35.2%) assigned to lower PEEP (with no significant difference between these two groups). Combining these data suggests a CFR in such patients with acute lung injury of 34.1% (783/2299). This estimate is a little less than one case series of ICU patients with acute respiratory failure from Influenza A (H1N1) and requiring mechanical ventilation, who had a CFR of 46% (156/337) [11]. Another study of such ICU patients with influenza found a lower overall CFR of 26% (492/1859) [12] and another one reported a CFR of 24% (177/733) [13]. Furthermore, in a group of 340 ICU patients with ARDS, the CFR at 90 days was 32% in a group given a neuromuscular blocker and 41% in the placebo group [14].

We then applied the 34.1% value from the meta-analysis to the data in Table 1, giving an estimated CFR of 1.37% ([14 x 34.1%]/349) for COVID-19. The 95% confidence interval (CI) of this estimate is 0.57% to 3.22%. This estimate can be considered a symptomatic CFR (sCFR) as it likely to be based on a denominator who had symptoms and were tested and laboratory-confirmed as cases. This estimate is almost invariably higher than the infection-fatality risk (IFR) which can only be estimated when serological testing becomes available to identify all of those infected [15].

Decision-makers and disease modellers might still be best to assume that the ‘true’ CFR is less than our estimate here as mild cases may not be identified. Furthermore, if a specific treatment is identified in the near future, then cases could have improved survival and the CFR might decline. On the other hand, if health systems became overloaded with the COVID-19 epidemic, then the CFR could likely increase for both the community and in hospital cases. As there is further progression of the COVID-19 epidemic, ongoing work will be needed to better clarify both transmissibility and the CFR, so that the likely mortality burden can be estimated. The method we have used here for CFR estimation will only be valid prior to healthcare systems becoming overloaded, after which they are likely to change their ICU admission policies.

Our provisional CFR estimate for COVID-19 is less than that estimated for MERS-CoV and SARS-CoV, but is broadly comparable with two of the three previous influenza pandemics (when considering the range of estimated values) (Table 2).

**Table 2.** Case fatality risks for selected pandemics (by descending order of mid-range estimates of magnitude)

| **Pandemic** | **CFR** | **Comments and references** |
| --- | --- | --- |
| Middle East respiratory syndrome coronavirus (MERS-CoV) | 34.4% | WHO estimate [16]. |
| Severe acute respiratory syndrome coronavirus (SARS-CoV) | 9.6% | Based on 8,096 reported cases, including 774 deaths in 27 countries (by July 2003) [17]. This could however be an over-estimate given the possibility of mild cases not being detected. |
| Pandemic influenza in 1918-1919 (United States [US]) | 0.5% to 5.3% depending on age-group | Highest in those aged <1 year (at 5.2%) and in the very old (70 to 74 years, 5.3%) [18]. Lowest in young children (5 to 9 years, 0.5%) and middle-aged adults (55 to 59 years, 0.9%). There was an intermediate peak in the CFR among young adults (25 to 29 years, 3.1%). |
| COVID-19 outside of China (this study) | 1.5% (0.6% to 3.5%) | These values could, however, be over-estimates given the possibility of mild cases not being detected (see main text). |
| Pandemic influenza in 2009 | 0% to 1.2% (but mainly 0.05% to 0.005%) | A systematic review considered 77 estimates from 50 published studies [19]. The values for symptomatic cases had point estimates ranging from 0 to 1,200 per 100,000 cases. Nevertheless, “most of the estimates in this category [symptomatic cases] fell in the range of 5 to 50 deaths per 100,000 cases.” |
| Pandemic influenza in 1957 (US data) | 0.2% | Calculated using US data from: [20]. |

## Data Availability

All data are displayed in the article as it is.

